# A Cross sectional Feasibility Study to Evaluate the Usability and Efficacy of Swaasa AI Platform for Rapid Respiratory Health Assessment

**DOI:** 10.1101/2023.10.02.23296452

**Authors:** Siva Kumar Lotheti, Haritha Vissamsetti, kiran Pamarthi, Devi Madhavi Bhimarasetty, Gowrisree Rudraraju, Narayana Rao Sripada, Charishma Gottipulla, Priyanka Firmal, Baswaraj Mamidgi, Shubha Deepti Palreddy, Nikhil kumar Reddy Bhoge, Harsha Vardhan Reddy Narreddy, Venkat Yechuri, Manmohan Jain, Venkata Sudhakar Peddireddi, Niranjan Joshi, Shibu Vijayan, Sanchit Turaga, Vardhan Avasarala

## Abstract

Analysing cough sounds is vital in pulmonary medicine. Recently, AI tools are being trained to analyse the acoustic signals of cough sounds so that more cases can be quickly tested, thereby reducing the patient load on primary healthcare systems. In this paper, we evaluate ”Swaasa”, our AI-based platform for rapid respiratory screening, highlighting its efficacy and ease of use. We applied our trained classifier to catch underlying pathologies from cough sound data collected from diverse sources. We then used a pattern classifier to identify specific respiratory conditions based on cough sound patterns. We tested the robustness of our methods by comparing our results with that of a pulmonary physician in 355 cases and show that Swaasa correctly predicted associated risk in 87.32% of those cases. Our platform has a sensitivity of 97.27% with a Positive Predictive Value (PPV) of 88.54%, giving us the potential to revolutionise disease screening, especially for large populations and in isolated rural areas. Our rapid and easy-to-use Software as a Service (SaaS) solution efficiently diagnoses and conserves resources, improves patient outcomes, and establishes a comprehensive and accessible healthcare framework.

## Introduction

A cough is a reflex action that clears the respiratory passages of excessive mucus, foreign particles, or other irritants [1]. It is a common symptom of many respiratory conditions, including the common cold, influenza, pneumonia, asthma, and chronic obstructive pulmonary disease (COPD) [2, 3]. Coughs can vary in duration, frequency, and intensity, and these variations can provide important information about the underlying condition [4]. Different types of coughs can be linked to different respiratory conditions. For example, a dry, hacking cough may be a symptom of a viral infection such as the common cold, while a wet cough that produces a lot of mucus may be a symptom of a bacterial infection or COPD. Coughs that are accompanied by wheezing or shortness of breath may indicate asthma [5].

Medical diagnostics of respiratory pathology involve a combination of history, examination using a stethoscope, spirometry or pulmonary function testing and imaging including chest X-ray, CT of the thorax and bronchoscopy. However, this is a resource intensive process which relies on appropriate infrastructure that may not be present in resource limited settings - this can prove difficult in diagnosing respiratory diseases [7, 8]. Hence there is a need for inexpensive and accessible remote pre-screening tools that can quickly and accurately identify cases of respiratory disorders, particularly in low- and middle-income countries where resources are limited. A remote screening tool could include portable diagnostic devices, telemedicine, or mobile health (mHealth) applications that can be used in the community or at the point-of-care [13, 14].

In the era of Artificial intelligence (AI), the use of cough sound analysis as a pre-screening tool for respiratory health has gained significant attention [15]. AI-powered algorithms can be trained on large datasets of cough sounds to identify patterns and characteristics associated with specific respiratory conditions, such as tuberculosis (TB), COVID-19, asthma, and COPD [16–18]. These algorithms can be integrated into portable diagnostic devices or mobile health (mHealth) applications, allowing for real-time screening of respiratory disorders in the community or at the point-of-care. There is a significant amount of research on using the characteristics of cough sounds for objective clinical assessment. A recently conducted double-blind, prospective, diagnostic accuracy study evaluated the performance of the algorithm by comparing it to expert clinical opinion and standard lung function testing. The accuracy of the algorithm in identifying asthma exacerbations, as measured by positive percent agreement with expert clinical diagnosis was 89% [19]. There are commercial devices available, such as the VitaloJAK Cough monitoring device and NuvoAir, which objectively measure cough by counting cough events automatically. These devices can be used to aid in the diagnosis of various diseases, including COPD and asthma [20, 21]. However, the main drawback of both these devices is the small sample size, which can limit the accuracy of the model. Additionally, relying on open-source data can also be a limitation, as the data may not be representative of the population or diverse enough to generalize to different patients. Although AI-based analysis of cough sounds is an innovative approach that has the potential to be an effective mass pre-screening tool for respiratory diseases, there remains a need for further research to validate the findings and improve the performance of the models.

In this study, we employed the "Swaasa" AI platform to analyse cough sounds and ascertain the presence or absence of underlying disease conditions, categorized as "Risk Yes" or "Risk No." Additionally, we investigated cough sound patterns to identify specific respiratory conditions, including normal patterns as well as those indicative of obstructive, restrictive, and mixed pathologies. The platform underwent training using data obtained from diverse subjects in multiple previous clinical trials, ensuring a substantial level of reliability and accuracy. Stringent measures were implemented to ensure data quality, resulting in a dataset devoid of noise and encompassing cough recordings from a diverse group of individuals. In contrast to conventional approaches, our methodology employed two parallel models. The first model utilized a Convolutional Neural Network (CNN) trained on Mel-frequency cepstrum (MFCC) spectrograms of cough sounds. The second model utilized primary and secondary features in a Feed-forward Artificial Neural Network (FFANN). This dual-model approach allowed for comprehensive analysis, leveraging the spectral information captured by the CNN model and the wider range of primary and secondary features used by the FFANN model. To further optimize the analysis, the final layers of these two models were merged, enabling the integration of their outputs and providing a more thorough and accurate assessment of the cough data. This fusion of models (combined logic) enhances the overall prediction capability of the platform [22]. To evaluate its effectiveness and feasibility a cross-sectional pilot study was conducted on 355 subjects at Simhachalam Rural Healthcare Centre (RHC). Upon comparing the results obtained from the Swaasa platform with the physician reports, the model had a sensitivity of 93.88% and a specificity of 75.48% when identifying underlying pathological conditions. Additionally, it was found to be 87.32% accurate in predicting the associated risk when compared to physician reports. Furthermore, there was a significant concordance between the respiratory disease pattern predicted by pulmonologists and the outcomes provided by Swaasa. The interrater variability, quantified by the Cohen’s kappa coefficient (κ=0.607), indicated a notable level of agreement among the raters.

The results obtained from Swaasa platform are highly promising, with the potential for it to be used for quick, cost-effective, and non-invasive pre-screening of respiratory health on a large scale. Further research is needed to make it more reliable for screening diverse subjects in different environmental conditions.

## Materials and Methods

### Data Collection

The cough data was collected at multiple sites, including Andhra Medical College, Visakhapatnam, India. The studies were registered under Clinical Trials Registry-India (CTRI/2021/07/035096), (CTRI/2021/09/036489) and (CTRI/2021/09/036609) were begun after getting the approval from the AMC-Institutional Ethics Committee (IEC) and all research was performed in accordance with relevant guidelines/regulations. The validation study cohort consisted of male and female patients above 18 years of age who presented with cough symptoms, assessed through a subset of the St. George’s Respiratory Questionnaire (SGRQ). Patients with suggestive symptoms of underlying respiratory conditions were included in the study. Informed consent was obtained from all participants and/or their legal guardians. To evaluate the model, patients’ demographic details and vital signs were collected, followed by interviews using the SGRQ questionnaire to gather information on respiratory symptoms and lung health. Trained healthcare personnel recorded cough sounds using smartphones, ensuring standardized procedures such as maintaining distance, angle, and duration. The collected cough sound samples were then subjected to noise filtering techniques, tailored to the device type. For smartphones with multiple microphones, a noise reduction algorithm based on power level ratios was applied, resulting in denoised audio recordings. For smartphones with a single microphone, noise removal techniques during pre-processing were employed to isolate the cough signal using Fast Fourier Transform. Valid coughs were detected using a cough/non-cough classifier, ensuring the inclusion of reliable data in the analysis. Safety measures were implemented during the recording process, including the use of surgical masks by subjects and sanitization of recording devices after each use. Additionally, reference standard tests, namely spirometry and chest X-ray examinations, were conducted to establish a clinical diagnosis. By comparing the model’s results with the clinical diagnosis based on the reference standard tests, the model’s performance in accurately assessing lung health and identifying specific respiratory conditions was evaluated.

### Model building

The collected cough data has been used to validate the feasibility and effectiveness of the model in screening respiratory conditions. The model’s architecture and functionalities were built as the research progressed. The architecture model is illustrated in figure 1. The audio recordings underwent rigorous processing and extraction procedures to isolate individual cough events. This involved careful removal of confounding factors, such as speech, background noise, and other respiratory sounds, to ensure the purity of the cough data. For the development of the Cough non-cough classifier, a dataset comprising cough sequences and non-cough sequences from a diverse population was utilized. The classifier was trained using machine learning techniques to differentiate between cough and non-cough sounds accurately. This dataset was carefully curated to encompass various scenarios encountered in real-world settings. The risk classifier, responsible for determining the presence of underlying respiratory problems, was trained on a separate dataset. It consisted of cough sequences labelled as Risk ’Yes’ (indicative of obstructive, restrictive, mixed, and other pathologies) and cough sequences labelled as Risk ’No’ (representing normal conditions). This dataset allowed the classifier to learn patterns and characteristics in the cough sounds associated with specific respiratory conditions. To identify respiratory disease patterns, a pattern classifier was developed using a combined logic approach. This classifier integrated a CNN and a FFANN model to automatically classify different types of respiratory diseases based on the signature patterns present in the cough data [22]. The classifier was trained on a dataset ensuring consistency and generalizability of the results. Throughout the development process, careful attention was given to the scientific rigor and reproducibility of the platform. The datasets used for training and validation were extensively validated and cross-validated to ensure the reliability of the classifiers. Additionally, the performance metrics of the classifiers were rigorously evaluated using appropriate statistical measures.

**Figure 1:** Architecture diagram of the Machine learning classifier

### Safety and Usability assessment

In the current study, we employed various methods to assess the usability and efficacy of the model for respiratory health assessment in a primary healthcare setting. The evaluation included Critical Task Analysis and a Questionnaire survey. Critical Task Analysis focused on analysing the user interface elements, while the Questionnaire survey aimed to assess user satisfaction. Our participant pool consisted of two health workers from a primary healthcare center and a qualified healthcare practitioner.

### Performance validation of model

Clinical validation was conducted to evaluate the effectiveness of the device when deployed as a screening tool prior to diagnosis. We enrolled 400 subjects from a peripheral health care centre, RHC Simhachalam. Figure 2 shows the age and gender distribution of the recruited subjects. Cough data from 45 subjects was inconclusive, hence considered data collected from 355 subjects only for the study.

**Figure 2:**
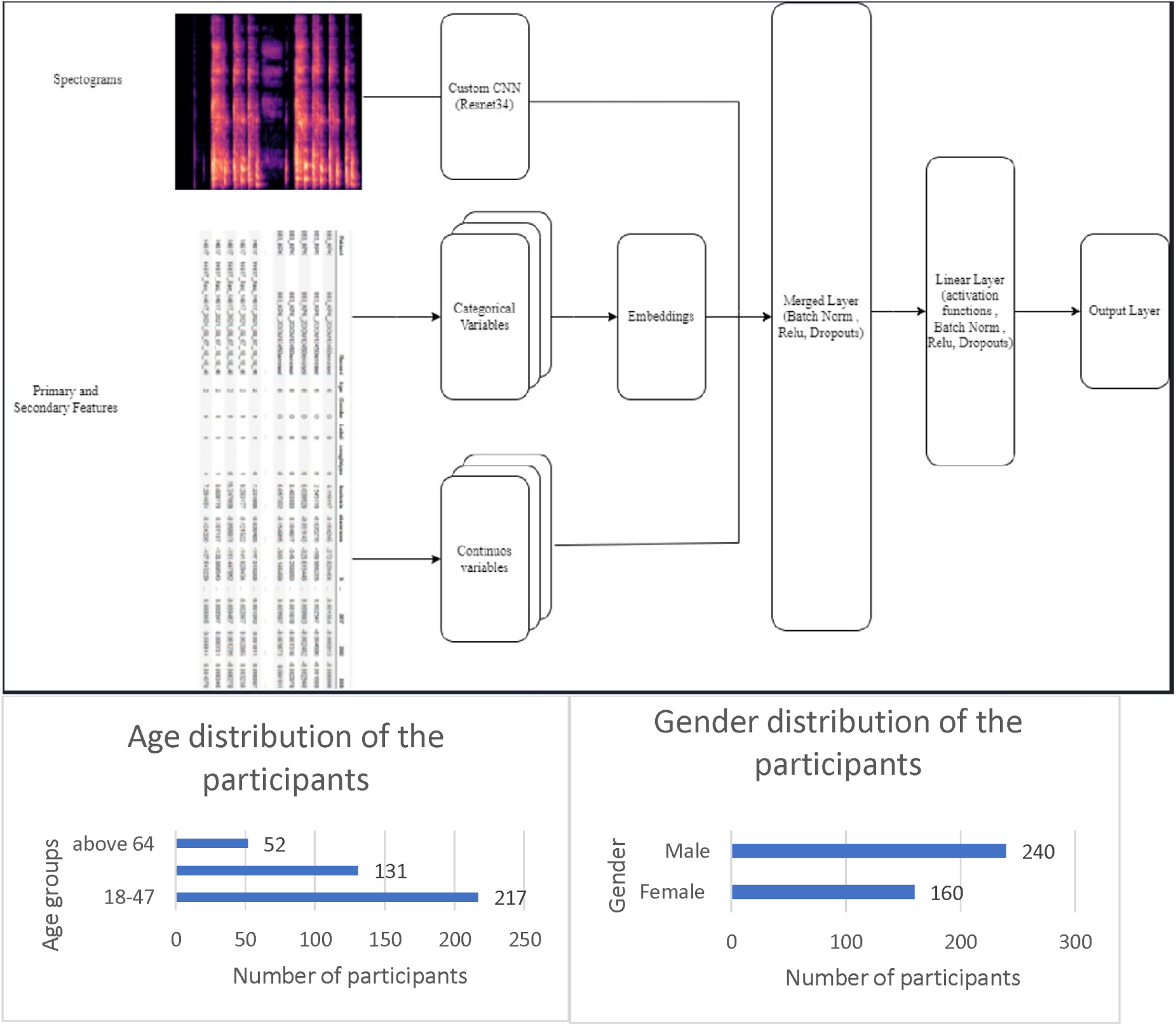
Data distribution in terms of age and gender in the Clinical Validation of the model

### Data analysis

A summary sheet was created by a statistician that includes the clinical diagnosis and the results from Swaasa platform. This sheet was used to compare the results against the gold standard, which is the diagnosis made by a physician. The physicians were not made aware of the results from the Swaasa system. The effectiveness of the model was determined by calculating the ratio of patients who were correctly diagnosed as positive to the total number of patients who had positive results by Swaasa.

### Statistical analysis

The comprehensive evaluation of the model performance on the test set includes accuracy sensitivity, specificity, positive prediction value (PPV), negative predictive values (NPV) and ROC. To measure the variability around these parameters, we used 95% confidence intervals using the Clopper–Pearson method [23]. We also used kappa (κ) statistic to test interrater reliability to better understand the performance of the model in predicting the respiratory disease pattern with respect to the Physician’s report.

## Results

### Model Output in the Clinical Validation

#### a) Risk Classifier

In the study, 355 individuals were screened, and the risk classifier successfully identified 286 individuals with potential respiratory disorders (Table 1). This resulted in an impressive positive predictive value of 88.54%. The ROC curve of risk classifier is in figure 3.

**Figure 3:**
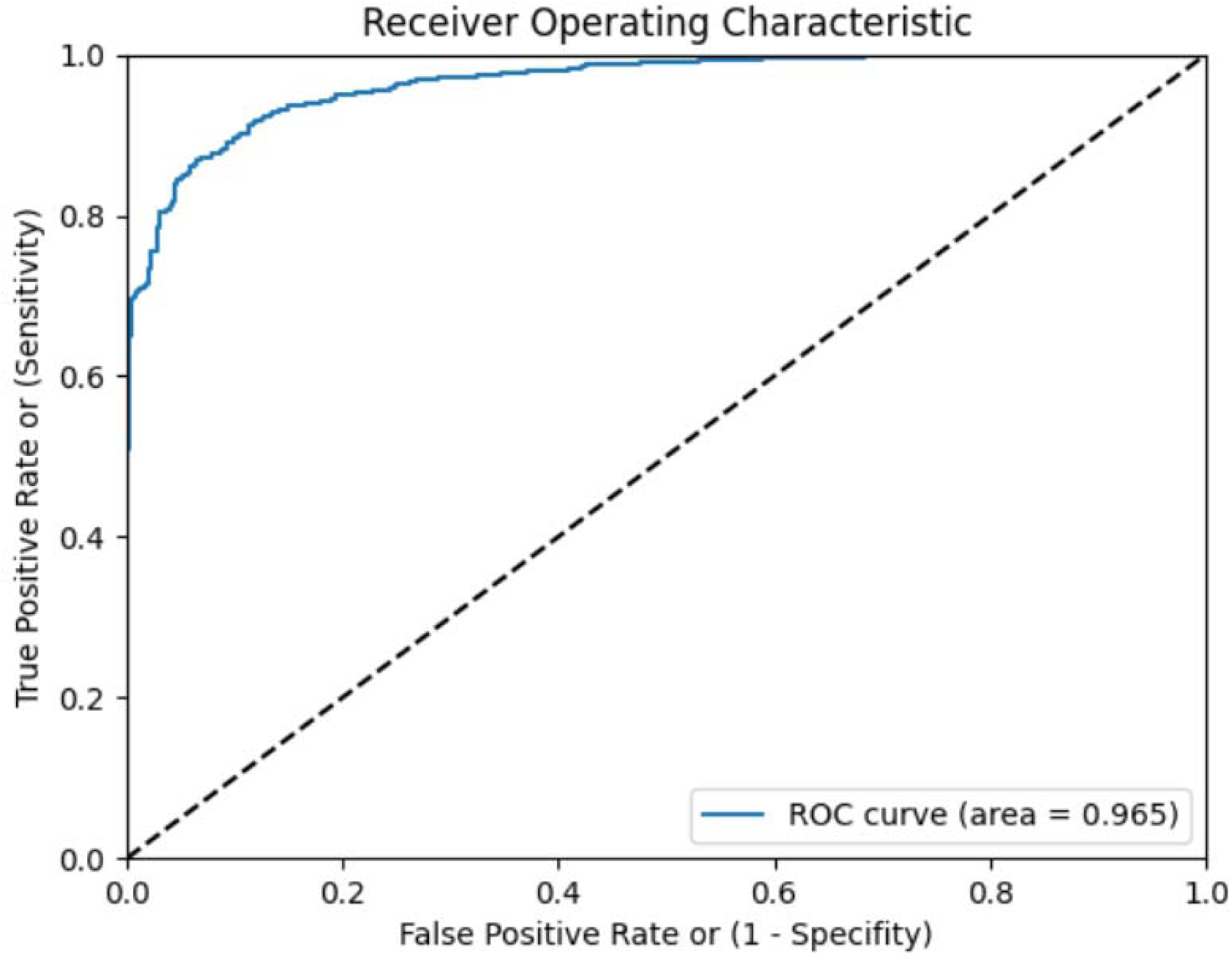
ROC curve for risk classifier

**Table 1:**
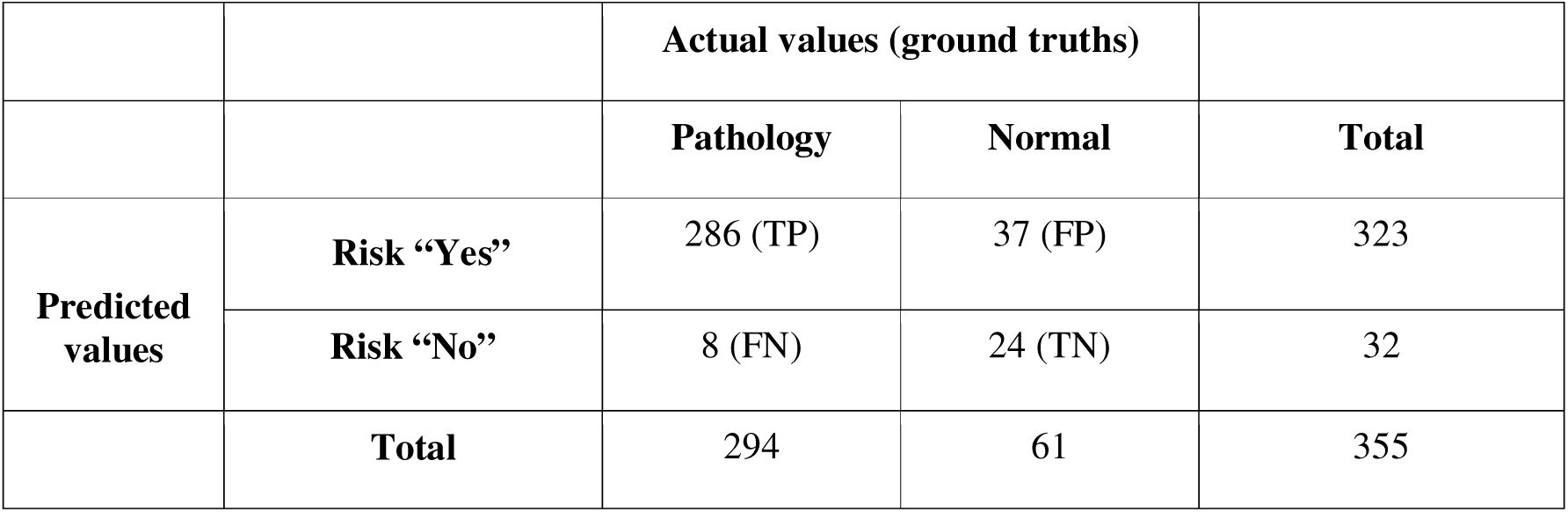
Confusion Matrix for the Risk Classifier depicting the classification outcomes of individuals as Risk “Yes" or Risk “No" for potential respiratory disorders.

Furthermore, the sensitivity of the risk classifier was determined to be 97.27%. This means that the system was able to accurately detect a large proportion of individuals with respiratory disorders, minimizing the chances of false negatives. On the other hand, the accuracy of the risk classifier was found to be 87.32% (Table 2). While not as high as the sensitivity, the accuracy still indicates that the system can reliably identify individuals without respiratory disorders, reducing false positives. The risk classifier’s accuracy and reliability make it an invaluable tool for early detection and intervention. Figure 4 illustrates the LIME (Local Interpretable Model-agnostic Explanations) maps, method for explaining the predictions of any machine learning model by approximating it with an interpretable model (such as a linear model) locally around the prediction. LIME provides a more general explanation of the model’s behaviour for a specific input. By identifying individuals at risk of respiratory disorders, healthcare professionals can initiate timely interventions, leading to improved patient outcomes. The high positive predictive value and sensitivity of the risk classifier make it an effective screening tool for identifying individuals who require further diagnostic testing and medical attention.

**Figure 4:**
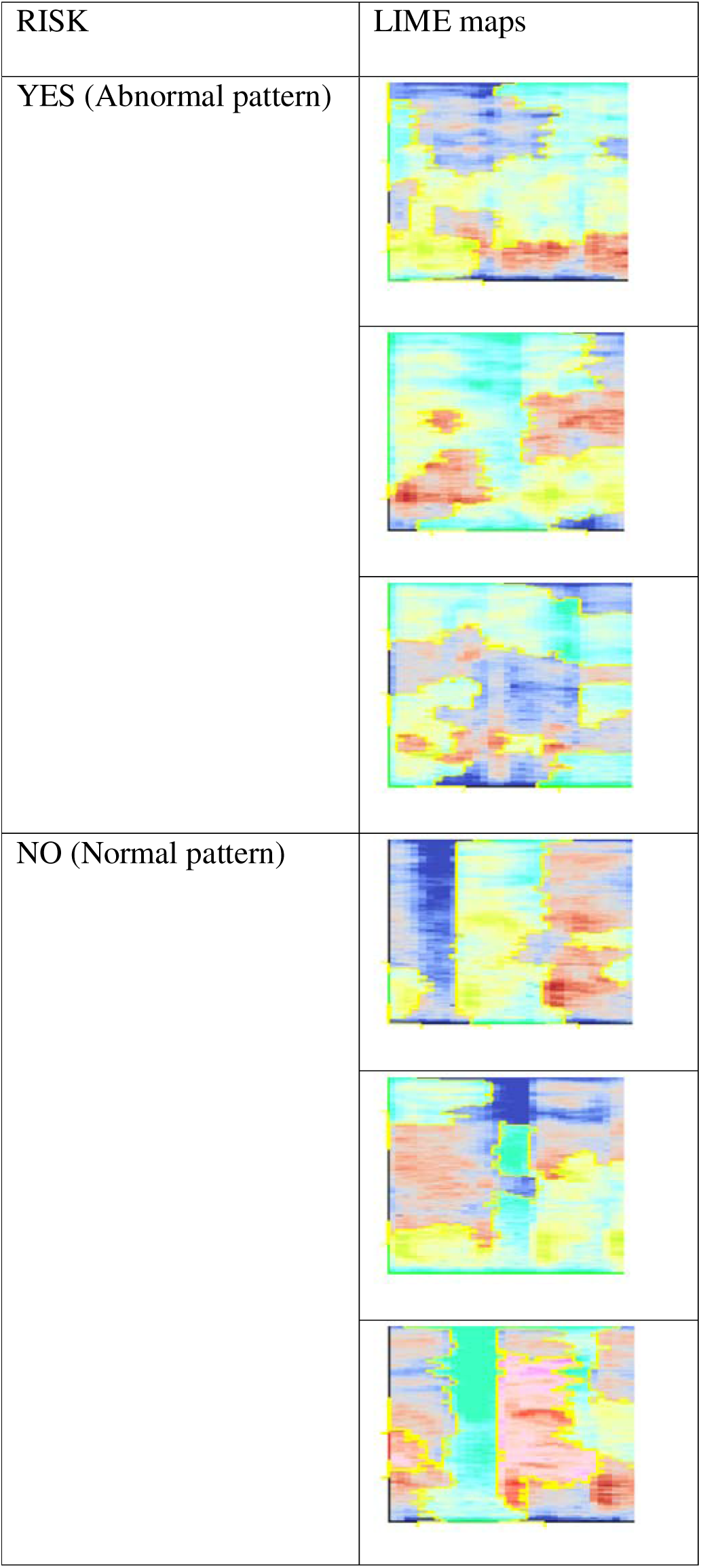
LIME maps of the coughs with normal and abnormal respiratory patterns.

**Table 2:**
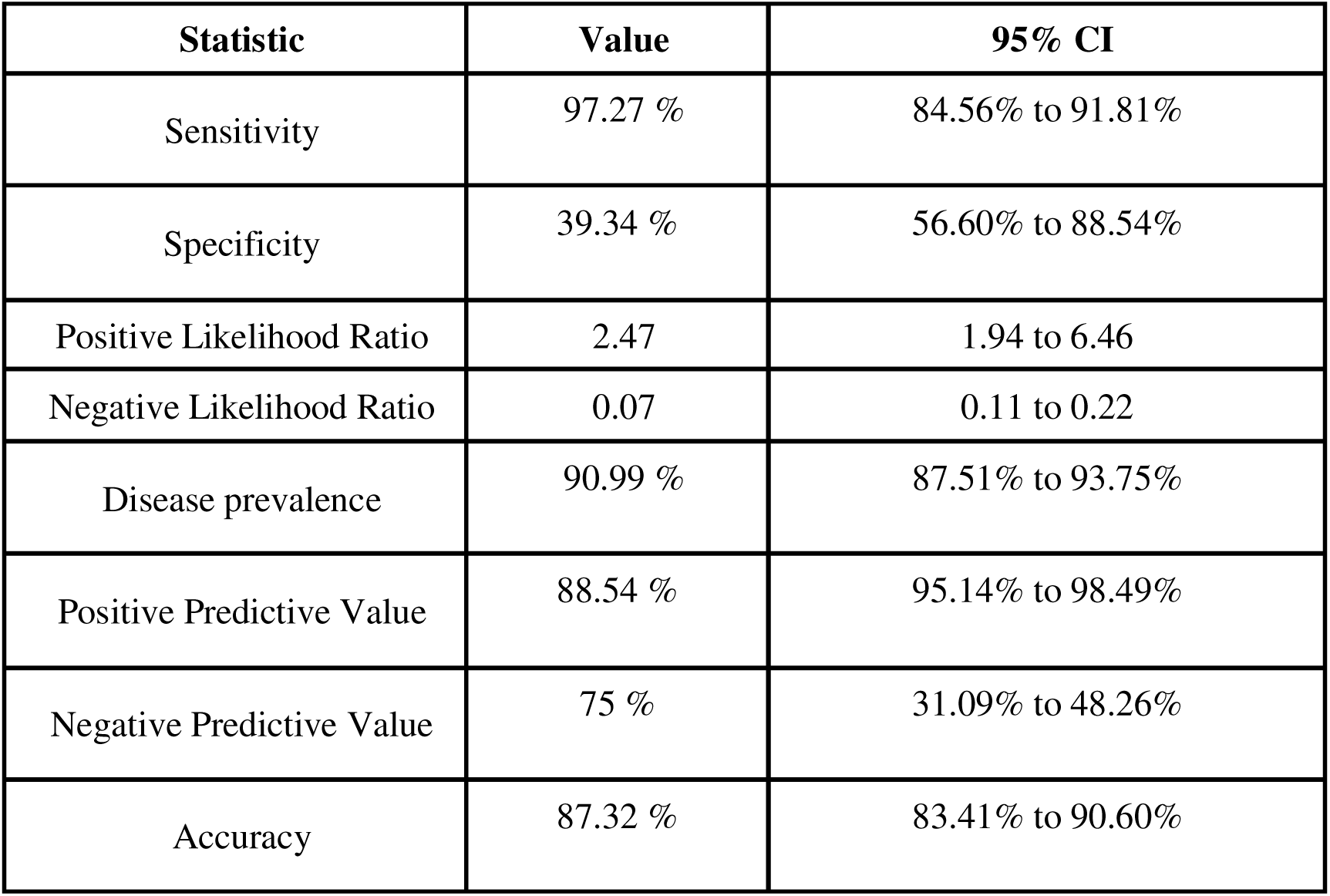
Performance metrics for the Risk classifier.

| Statistic | Value | 95% CI |
| --- | --- | --- |
| Sensitivity | 97.27 % | 84.56% to 91.81% |
| Specificity | 39.34 % | 56.60% to 88.54% |
| Positive Likelihood Ratio | 2.47 | 1.94 to 6.46 |
| Negative Likelihood Ratio | 0.07 | 0.11 to 0.22 |
| Disease prevalence | 90.99 % | 87.51% to 93.75% |
| Positive Predictive Value | 88.54 % | 95.14% to 98.49% |
| Negative Predictive Value | 75 % | 31.09% to 48.26% |
| Accuracy | 87.32 % | 83.41% to 90.60% |

#### b) Pattern Classifier

Model’s pattern classifier is designed to categorize individuals into different respiratory health patterns, providing valuable insights into their lung function and airflow patterns. The classifier successfully categorizes individuals into four distinct categories: "Normal," "Obstructive," "Restrictive," and "Mixed." A "Normal" classification indicates a healthy respiratory system with no significant abnormalities detected in lung function or airflow patterns. An "Obstructive" classification suggests the presence of airway blockage, which can be indicative of conditions such as asthma or chronic obstructive pulmonary disease (COPD). A "Restrictive" classification points to a decrease in the lung’s ability to expand fully, which may be caused by conditions like pulmonary fibrosis or chest wall deformities. Lastly, a "Mixed" classification suggests a combination of both obstructive and restrictive patterns (Table 3).

**Table 3:**
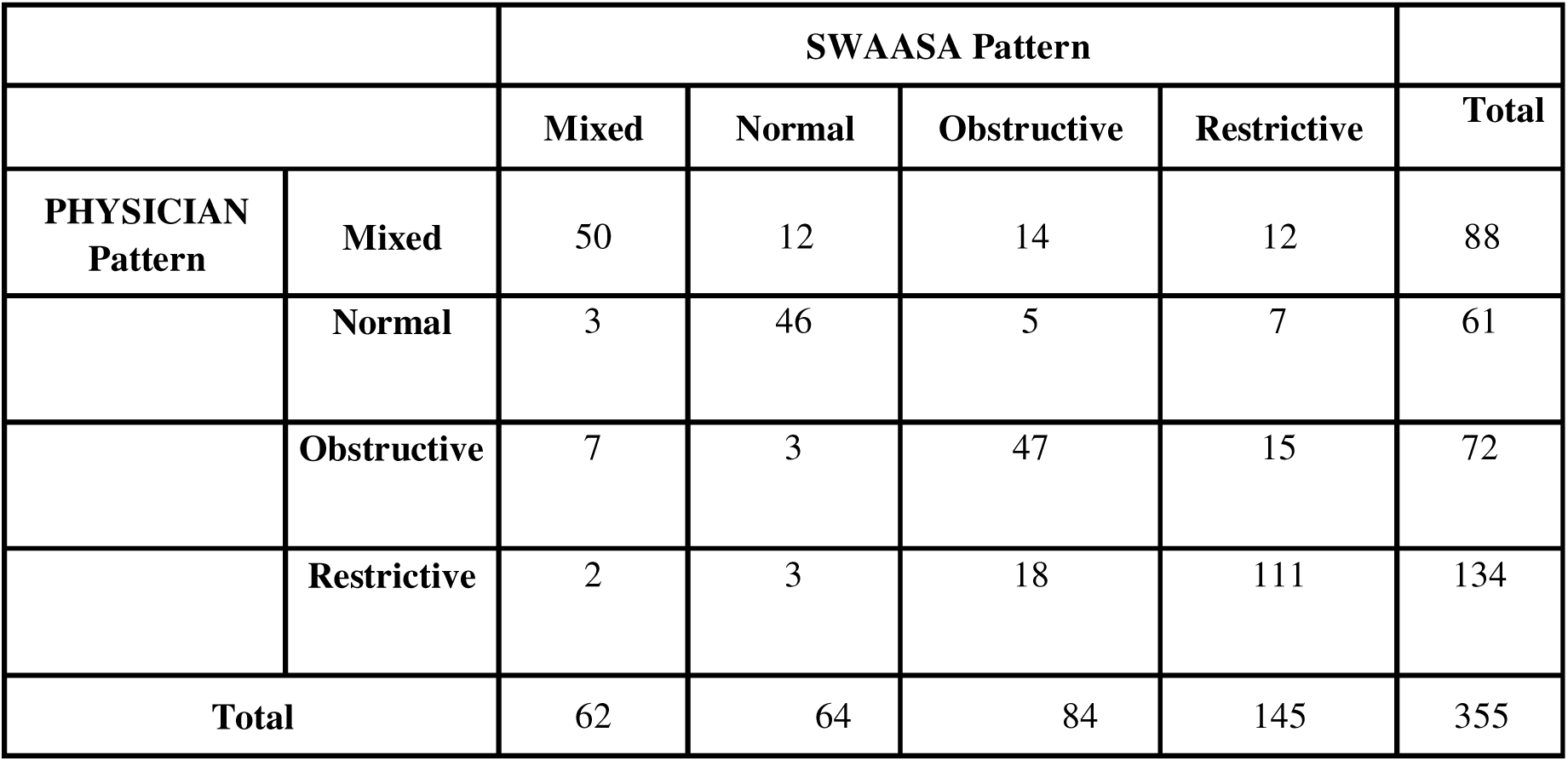
Table showing the comparison of different respiratory disease patterns obtained by Swaasa and Pulmonologist’s examination.

|  |  | Mixed | Normal | Obstructive | Restrictive | Total |
| --- | --- | --- | --- | --- | --- | --- |
| PHYSICIAN<br>Pattern | Mixed | 50 | 12 | 14 | 12 | 88 |
|  | Normal | 3 | 46 | 5 | 7 | 61 |
|  | Obstructive | 7 | 3 | 47 | 15 | 72 |
|  | Restrictive | 2 | 3 | 18 | 111 | 134 |
| Total |  | 62 | 64 | 84 | 145 | 355 |

To validate the accuracy and consistency of the pattern classifier, a comparison was made between the classifier’s predictions and the assessments made by pulmonologists. The study found a moderate level of agreement (κ = 0.607) between the predictions of the pulmonologists and the outcomes generated by the Swaasa system (Table 4). This level of agreement signifies that the pattern classifier’s categorizations align closely with the assessments made by experts in the field, further validating the system’s accuracy and reliability.

**Table 4:** kappa statistic to test interrater reliability of the respiratory disease pattern predicted by pulmonologist w.r.t. the Swaasa outcome.

| Symmetric Measures |  |  |  |  |  |
| --- | --- | --- | --- | --- | --- |
|  |  | Value | Asymptotic<br>Standard Error | Approximate T | P VALUE |
| Measure of<br>Agreement | Kappa ($\kappa$) | .607 | .033 | 19.423 | .001 |
| N of Valid Cases |  | 355 |  |  |  |

The pattern classifier’s ability to categorize individuals into different respiratory health patterns allows for targeted treatment and management plans. By understanding the specific pattern exhibited by an individual, healthcare professionals can tailor interventions to address the underlying condition effectively. This personalized approach improves the quality of care and outcomes for patients with respiratory disorders. Overall, Model’s pattern classifier provides valuable insights into an individual’s respiratory health, categorizing them into distinct patterns that aid in diagnosis and treatment planning. The agreement between the system’s predictions and those of pulmonologists validates its accuracy, while its simplicity and accessibility make it a practical tool for healthcare providers to incorporate into their respiratory health screenings.

### Model feasibility study

The evaluation of the model yielded important results regarding safety and usability. In terms of safety, potential hazards and failure modes associated with the platform were identified, such as the absence of standard cough recording protocols, adverse effects for specific patient conditions, background noise during audio recording, and inadequate precautions for device cleaning. Control measures were implemented, and safety guidelines were followed to mitigate these risks. Regarding usability, the Critical Task Analysis demonstrated that users were generally able to complete critical tasks successfully. However, some issues related to screen navigation and session expiry were identified. The Questionnaire survey indicated that users found the device interface pleasant, easy to learn, and consistent. Areas for improvement were identified, such as expanding the coverage of error messages in the instructions for use (IFU) and addressing occasional app stability issues. Based on the study findings, several suggestions for improvement were compiled, including enhancing screen navigation, addressing session expiry issues, expanding the coverage of error messages in the IFU, and resolving occasional app stability issues. Overall, the participants expressed satisfaction with the simplicity and ease of use of the device. In conclusion, our feasibility study confirms the usability and safety of the Swaasa AI platform for respiratory health assessment in a primary healthcare setting. The study findings have provided valuable insights and recommendations for further enhancing the device’s usability and effectiveness.

## Discussion

Machine learning (ML) has the potential to revolutionize the way respiratory diseases are detected and diagnosed. By combining cough sound analysis with computed tomography (CT) and chest radiography (CXR), ML algorithms can efficiently detect a wide range of conditions, including COVID-19 [24, 25].

Such methods have already been used in the past to diagnose and predict the outcome of conditions such as lung cancer, bronchitis, pneumonia, COPD, TB and asthma [5, 17, 26–31]. AI has the potential to improve respiratory health by providing faster and more accurate diagnoses. AI algorithms can analyse large data sets to identify patterns and trends that help in developing new treatment methods. Our contribution to the State of the art is the Swaasa AI platform that combines the final output layers of two separate models - a tabular model that uses the primary and secondary features as input and a CNN model that used MFCC spectrograms as input. This approach resulted in significantly better predictions than previously used logistic regression or a CNN model alone [17, 30, 31].

In our cross-sectional feasibility study, we evaluated the usability and efficacy of "Swaasa" for rapid respiratory health assessment. The study specifically focused on analysing cough patterns and using risk classifiers to assess respiratory health. The risk classifier, trained using cough data, accurately determines the risk of respiratory disorders, classifying it as "Yes" or "No." This allows for early identification of individuals who may require further evaluation and medical intervention. The pattern classifier, also trained on cough data, categorizes coughs into three types: obstructive, restrictive, and mixed, providing valuable insights into the underlying respiratory conditions. Together, these classifiers enable a comprehensive assessment of respiratory health, facilitating timely interventions and personalized treatment plans. Also, the clinical validation was conducted on a relatively large cohort, whereas previous studies were carried out on smaller scales or using unreliable crowdsource datasets [17, 30, 32]. This approach provided a more robust and reliable dataset for evaluation.

Model’s risk classifier plays a crucial role in identifying individuals at risk of respiratory disorders. By analysing cough patterns, the platform accurately determines the likelihood of respiratory issues, such as obstructive, restrictive, and mixed conditions. The study revealed that the risk classifier of the Swaasa platform achieved a sensitivity of 97.27%, indicating its high capability in correctly identifying individuals who are at risk. This sensitivity implies that the risk classifier has a low rate of false negatives, effectively detecting a significant proportion of individuals with potential respiratory disorders. The accurate identification of at-risk individuals enables timely interventions and targeted healthcare strategies to improve their overall respiratory health outcomes. The results of the assessments obtained using the model were found to be highly correlated with those obtained from traditional methods, indicating that the platform can accurately assess respiratory health. The results of the study also showed that the predictions made by pulmonologists regarding respiratory disease patterns matched well with the outcomes generated by the Swaasa system. This was demonstrated by a moderate level of agreement, as measured by the interrater variability value (κ = 0.607). This indicates that the predictions made by pulmonologists and the outcomes generated by Swaasa system were consistent, which is important validation of the accuracy and reliability of the system.

The present study establishes the feasibility and safety of implementing the Swaasa AI platform for respiratory health assessment in primary healthcare settings. Through a comprehensive evaluation involving Critical Task Analysis and a Questionnaire survey, valuable insights were gained regarding the platform’s usability and efficacy. The study identified important safety considerations, including the need for standardized cough recording protocols, mitigation of potential adverse effects, management of background noise during audio recording, and implementation of appropriate device cleaning procedures. Usability enhancements were recommended, particularly in addressing screen navigation and session expiry issues. Additionally, the study highlighted the high level of user satisfaction, with participants expressing positive feedback regarding the platform’s simplicity, ease of use, and consistency. These findings reinforce the potential of the Swaasa AI platform as a valuable tool for respiratory health assessment in primary healthcare, while also providing valuable guidance for further refinement and optimization.

In conclusion, the model demonstrates the feasibility and efficacy of utilizing cough patterns and risk classifiers for rapid respiratory health assessment. Its potential applications extend to primary care clinics, hospitals, and remote locations, where it can significantly enhance the speed and accuracy of respiratory health evaluations. Furthermore, the platform serves as a valuable triage tool, enabling the identification of high-risk patients and optimizing the allocation of clinical resources for improved patient outcomes.

To ensure the generalizability of the Swaasa platform, future studies will involve mass screening of subjects from diverse populations, languages, and cultures. This approach will validate its effectiveness and applicability across various settings globally. Additionally, efforts will be made to diversify the training data set, incorporating a wider range of respiratory conditions and demographics, further enhancing the platform’s accuracy and reliability.

Safety and usability considerations are integral to the continuous development and implementation of the model. Strict adherence to safety guidelines, including the standardization of cough recording protocols, mitigation of potential adverse effects, management of background noise, and appropriate device cleaning procedures, ensures the safety of both patients and healthcare providers. Ongoing refinements will address usability aspects such as screen navigation and session expiry issues, aiming to optimize user experience and facilitate seamless integration of the platform into routine clinical practice.

Overall, the model demonstrates promise as an efficient and reliable screening tool for respiratory health assessment, with future advancements and validations anticipated to further establish its utility in diverse healthcare settings.

## Data Availability

All data produced in the present study are available upon reasonable request to the authors.

## Data availability

Due to the nature of this research, participants of this study did not agree for their data to be shared publicly. However, the detailed analysis can be shared by NRS upon reasonable request.

## Author contributions

SKL and DMB defined study protocol, including the study design and methodology. NR conceptualized the idea of using cough sounds for screening and diagnosing respiratory health. GR performed literature review. BM, NKRB, HVRN, SDP, and CG were involved in device development. VY and MJ created value proposition for the device. HV assisted in executing the project at AMC by providing all the resources and extending research capabilities. GR, CG and KP performed data analysis, sample size estimation and result analysis. VSP, NJ, SV, ST, VA provided subject matter expertise. GR and PF wrote the manuscript. All the authors provided intellectual inputs and helped in preparing the manuscript.

## Acknowledgement

This study is supported by the MietY (Ministry of Electronics and Information Technology), India. We would also like to acknowledge the team from Andhra Medical College Visakhapatnam for all the support provided.

